# Relating SARS-CoV-2 shedding rate in wastewater to daily positive tests data: A consistent model based approach

**DOI:** 10.1101/2021.07.04.21259903

**Authors:** M. Petala, M. Kostoglou, Th. Karapantsios, Ch. Dovas, Th. Lytras, D. Paraskevis, E. Roilides, A. Koutsolioutsou-Benaki, G. Panagiotakopoulos, V. Sypsa, S. Metallidis, A. Papa, E. Stylianidis, A. Papadopoulos, S. Tsiodras, N. Papaioannou

## Abstract

During the COVID-19 pandemic, wastewater-based epidemiology (WBE) has been engaged to complement medical surveillance and in some cases to also act as an early diagnosis indicator of viral spreading in the community. Most efforts worldwide by the scientific community and commercial companies focus on the formulation of protocols for SARS CoV-2 analysis in wastewater and approaches addressing the quantitative relationship between WBE and medical surveillance are lacking. In the present study, a mathematical model is developed which uses as input the number of daily positive medical tests together with the highly non-linear shedding rate curve of individuals to estimate the evolution of virus shedding rate in wastewater along calendar days. A comprehensive parametric study by the model using as input actual medical surveillance and WBE data for the city of Thessaloniki (∼700,000 inhabitants, North Greece) during the outbreak of November 2020 reveals the conditions under which WBE can be used as an early warning tool for predicting pandemic outbreaks. It is shown that early warning capacity is different along the days of an outbreak and depends strongly on the number of days apart between the day of maximum shedding rate of infected individuals in their disease cycle and the day of their medical testing. The present data indicate for Thessaloniki an average early warning capacity of around 2 days. Moreover, the data imply that there exists a proportion between unreported cases (asymptomatic persons with mild symptoms that do not seek medical advice) and reported cases. The proportion increases with the number of reported cases. The early detection capacity of WBE improves substantially in the presence of an increasing number of unreported cases. For Thessaloniki at the peak of the pandemic in mid-November 2020, the number of unreported cases reached a maximum around 4 times the number of reported cases.

**HIGHLIGHTS:**

- Model estimates viral load evolution in wastewater from infected people dynamics
- Identifying actual conditions for which WBE can be used as an early warning tool
- Early warning capacity increases with an increasing number of unreported cases
- In Thessaloniki Nov20 outbreak, the early warning capacity of WBE was about 2 days
- In Thessaloniki Nov20 outbreak, unreported cases were up to 4 times reported cases

## INTRODUCTION

In the COVID-19 pandemic, wastewater-based epidemiology (WBE) has become an important tool, supplementing public health surveillance, in the hands of scientists, medical experts, and state officials to evaluate the spread of SARS-CoV-2 in the community (la Rosa et al., 2020; Medema et al., 2020b; Wurtzer1 et al., n.d.). This was more so after the Centers for Disease Control and Prevention (CDC) adopted wastewater disease surveillance as a valid monitoring tool and provided guidance and recommendations for the selection and application of testing methods (CDC, 2021). The latter cover issues from sample collection and sample processing to RNA measurement and use of laboratory controls for the estimation of the performance of the applied methods and of data quality. Laboratory controls deal with difficulties arising from the chemical and biological variability of wastewater quality across different places and over time, e.g. due to season, weather, human activities etc., and also cope with problems of proper RNA extraction and quantification, elimination of inhibition and contamination of reagents.

The so-called matrix recovery controls suggested by CDC refer exclusively to the amount of virus lost during sample processing. However, it is well known that viruses strongly adsorb, and so get inaccessible (“lost”), in the pores of solid particles suspended in wastewater in sewerage networks (Sellaoui et al., 2020; Ye, 2018). Therefore, if WBE studies aim to quantify the virus shedding rate at the source (households) from analysis of samples taken at the entrance of wastewater treatment plants (WTP) then it is of paramount importance to appraise the amount of virus lost in sewerage networks. Determining the extent of recovery, as suggested by CDC, by spiking techniques in samples obtained at the entrance of WTP is incapable of describing the amount of virus lost in sewerage networks. This is because such samples have traveled in sewerage pipes for hours and so have (most of) their adsorption sites saturated with adsorbed species. Desorption of spiked material would have been completely different if a sample had been taken fresh at a shedding spot, i.e., from the sanitary plumbing system of a building.

Recently, a mathematical model has been proposed to account for virus loss in sewerage networks by means of adsorption of virus on porous particles suspended in wastewater(Petala et al., 2021). To do so, the model rationalizes the SARS-CoV-2 concentration in samples taken at the entrance of WTP with respect to certain quality characteristics of wastewater samples. Rationalization is based on rigorous physicochemical phenomena in adsorption (and not on a statistical dependence of virus concentration on wastewater parameters), also including the effect of large-scale topological complexity of actual sewage networks. At the examined period of time (April to June 2020), the rationalized decreasing shedding rate was in agreement with the observed clinical conditions, contrary to the non-rationalized data which showed a different picture. Yet, even with rationalized data it is still difficult to make the critical step ahead and associate the virus shedding rate with the number of cases.

There are several reasons for the discrepancy between viral shedding rate in wastewater and number of infected people (cases) reported by public health surveillance. Limitations in the capacity of medical testing, strict criteria for the application of medical testing and parts of the population being reluctant to seek medical care are responsible for underdiagnosis of cases, mainly asymptomatic cases or patients with mild symptoms who are not tested. These factors lead to reporting a lower number of cases than the actual one. On the other hand, poor recovery in the collected wastewater samples and virus loss in sewage networks, along with ineffectiveness to properly rationalize for the latter, lead to a smaller measured concentration of virus in the collected samples and, thus, to a lower estimated viral shedding rate than the actual one.

The time delay between the onset of patients’ viral shedding in infected people and symptoms onset that prompts patients being tested, is another reason for discrepancy. Several reports in literature indicate a time delay up to 8 days between wastewater signal and infected cases reported in public health surveillance system (e.g.(D’Aoust et al., 2021; Nemudryi et al., 2020; Peccia et al., 2020)). This is in line with the reported incubation period of around 6 days from exposure/infection to onset of symptoms of COVID-19 (Guan et al., 2020; Lauer et al., 2020; Li et al., 2020). Therefore, the discrepancy between the onset of viral shedding and the onset of symptoms is actually an advantage that might explain the early detection capacity of WBE.

Another important reason of discrepancy between daily reports of wastewater viral titers and daily reports of new confirmed cases is the cumulative character of viral shedding rate in wastewater during the disease. Several clinical studies 14–16 showed that viral shedding is not uniform during the disease, but there is a maximum shedding rate in the very acute phase of infection, probably even before respiratory symptoms appear, being followed by an exponential decline in subsequent days. If one considers that shedding lasts at least 3-4 weeks after the inception of symptoms 14,15. it is apparent that daily wastewater viral titers correspond more to the cumulative shedding of infected people rather than to the shedding of the daily new cases. It must be stressed here that the evidence from clinical studies is limited, and only for samples collected from hospitalized patients, thus after symptoms onset, so one should be extremely careful in estimating shedding rates at the first days right after infection.

If one aims to associate wastewater surveillance data with public health surveillance data, the different sources of bias between these data should be also taken into account. Laboratory practice shows that variation in wastewater data is considerably high from one day to the next because of the many experimental parameters that are involved, e.g., individual’s shedding rate (orders of magnitude variation among infected people(Zheng et al., 2020); sampling protocol, sample concentration techniques, RNA extraction methodology, normalization with respect to population indicators and flow rate, inhibition assessment in RNA recovery, RNA quantification, etc. On the other hand, public health surveillance is susceptible to a systematic bias because of limitations in the capacity of medical testing along with the unpredictable human behavior under stressful conditions. In principle, one should be concerned about the correctness of both wastewater viral measurements and clinical testing data.

The goal of this work is to setup a methodology for the consistent comparison between the number of infected people reported by clinical surveillance and SARS-CoV-2 RNA concentration in wastewater samples. To do so, a model is developed for the estimation of the evolution of virus shedding rate in a sewage system based on the daily positive medical tests. Apart from corrections based on the laboratory controls suggested by CDC, virus shedding rate data are further rationalized with respect to physicochemical parameters of wastewater to account for virus loss by adsorption to sewage solids (Petala et al., 2021). To increase accuracy, clinical testing data are based on the date of specimen collection and not on the reporting date. The model is first developed in a generalized continuous time regime and then it is transformed to its discrete counterpart dictated by the daily basis data.

## METHODS

### Sampling

Wastewater samples were collected at the exit of Thessaloniki’s (a city at North Greece) main sewerage pipe, right before the entrance at the wastewater treatment plant of the city, as described elsewhere(Petala et al., 2021). This plant serves an estimated population of about 700,000 people. The present work reports 24-hours composite samples (1L each) taken three times per week (Monday-Wednesday-Friday) from October 5^th^, 2020 until January 6^th^, 2021. This period includes the second wave burst of COVID-19 for Thessaloniki in November 2020 which was the worst ever in the whole country. Typical range of values of the physicochemical parameters of wastewater samples for the examined period are displayed in Table 1 (supplementary materials). These parameters were employed in the rationalization of the measured viral concentration with respect to quality characteristics of wastewater according to Petala et al. (Petala et al., 2021).

### Virus concentration

Upon wastewater collection, 200 mL of each wastewater sample was centrifuged at 4000xg for 30 min to remove particles and pH of the supernatant was adjusted to 4 using a solution of 2.0 M HCl. An aliquot of 40 mL supernatant was then passed through an electronegatively charged surface of 0.45 μm-Ø47 mm cellulose nitrate HA membrane ((HAWP04700; Merck Millipore Ltd., Tullagreen, Ireland). Filtration was performed using a magnetic funnel mounted on a glass filtration flask (Pall Corporation) Filtration step was conducted in triplicate and membranes were stored in 15 mL falcon tubes for further processing of RNA extraction and virus quantification.

### RNA extraction and virus quantification

For each sewage sample, three electronegative membranes were individually subjected to phenol-chloroform-based RNA extraction process (Chaintoutis et al., 2019) coupled with magnetic bead binding. Each membrane filter was rolled into a Falcon™ 15 mL conical centrifuge tube with the top side facing inward. The following were added sequentially, a) 900 μL of guanidinium isothiocyanate-based “Lysis buffer I” [5M guanidinium isothiocyanate, 25mM EDTA, 25mM sodium citrate (pH 7.0), 25mM phosphate buffer (pH 6.6)] containing 1% N-Lauroylsarcosine, 2% Triton X-100, 2% CTAB and 2% PVP, b) 18μl β-mercaptoethanol, c) 300μl H2O, mixed thoroughly by inversion, and each tube was incubated at 4 °C on horizontal rotator (50 rpm) for 10-30 min. 1200 μl of “Lysis buffer II” were added [prepared by mixing 152.5gr guanidinium hydrochloride, 31.25 ml of 2M acetate buffer (pH 3.8) and water-saturated phenol stabilized (pH4), up to 500ml final volume], followed by incubation 10 min / RT on horizontal rotator (150 rpm). The liquid phase was transferred on a 2-mL microcentrifuge tube and clarified by centrifugation (21,000×g, 5 min, 4 °C). 1600 μl were transferred to a new tube, 200 μl chloroform-isoamyl alcohol (24:1) were added and shaken vigorously for 30 s followed by incubation (−20 °C, 30 min) and centrifugation (21,000×g, 10 min, 4 °C). 800 μl of the upper aqueous phase were transferred and mixed with 667μl isopropanol and 20μl of magnetic beads (IDEXX Water DNA/RNA Magnetic Bead Kit), followed by incubation 15 min / RT on horizontal rotator (150 rpm). The beads were washed according to the manufacturer’s protocol, using the magnetic extraction reagents. Elution of RNA was done in 100 μl buffer followed by filtering using OneStep PCR inhibitor removal kit (Zymoresearch).

SARS-CoV-2 quantified RNA (1× 106.3 genomic copies) from human clinical samples was added to a subset of concentrates to estimate the recovery efficiency and reproducibility of the RNA extraction procedure. Similarly, heat inactivated SARS-CoV-2 from human clinical samples was spiked (1× 107 viral particles) in a subset of sewage samples to assess the recovery and reproducibility of the virus concentration procedure.

Each RNA extract was subjected to real-time RT-PCR for SARS-CoV-2 quantification in triplicates. Two primer/probe sets were utilized: the N2 set from CDC that targets the nucleocapsid (N) gene and the set targeting the genomic region that encodes the E protein (Corman et al., 2020). The assays were performed on a CFX96 Touch™ Real-Time PCR Detection System (Bio-Rad Laboratories, Hercules, CA, USA). Reactions were considered positive if the cycle threshold was below 40 cycles. Calibration curves were generated using the synthetic single-stranded RNA standard “EURM-019” (Joint Research Centre, European Commission). The possible presence of RT-PCR inhibitors in each RNA extract was assessed in duplicates using spiked EURM-019 included at ∼1000 copies in additional RT-PCR reactions. Inhibition was expressed as % reduction in reported copy number, compared to the sum of spiked EURM-019 copies and the mean value of measured SARS-CoV-2 genomic copies in the non-spiked RNA. SARS-CoV-2 viral load in each sewage sample was expressed as mean ± standard deviation genome copies per liter, after correcting for RT-PCR inhibition if present, and recovery efficiencies (virus concentration and RNA extraction). Precision of each individual sewage sample quantification was assessed using the coefficient of variation (CV) of the estimated SARS-CoV-2 viral load of the three electronegative membranes processed. We set our precision threshold at 35% CV for each sewage sample measurement.

### Epidemiological data

Daily numbers of COVID-19 infected people reported in the city of Thessaloniki, adjusted to specimen collection date, were obtained from the National Public Health Organization (National Public Health Organization, 2020). These data reflect residents found positive when tested in public and private laboratories. Data are presented for the period between September 1^st^, 2020 and January 6^th^, 2021 (original data are provided in supplementary materials).

### Problem formulation

A model is developed for estimating the evolution of virus shedding rate to sewage system based on the official (announced by the state) results of daily positive tests (cases), registered by the date of specimen collection and not by the date of public reporting. There is typically a delay of few days between specimen collection and public reporting. The model is first developed in a generalized continuous time regime and then it is transformed to its discrete counterpart dictated by the daily basis data.

Let us denote as f(t) (t: calendar day) the evolution of positive test counts density with f(t)dt being the number of positive tests in the time period between t and t+dt. The first step is to transform the function f(t) to the function F(t) which denotes the total number of reported infected people at time t. It is essential to stress here that F(t) includes people that at time t either had already a positive test or they are in the first days after infection, so do not have symptoms, but still they shed virus and will be tested positive later after their onset of symptoms. The effect of infected but unreported people, such as asymptomatic ones and those not tested because of mild symptoms, is dealt with later.

In order to proceed let us consider the course of the disease of an infected person: Infection starts (disease incidence) at day τ=1, detection occurs (specimen collection) at day τ=τ_d_ and end of viral shedding occurs at day τ_e_. The number of days for detection and end of shedding are in general not the same among cases but they show a dispersion which can be described by the respective probability density functions P_d_(τ_d_) and P_e_(τ_e_). It is understood that both functions take non-zero values only in restricted domains of their arguments. The limits of these domains are denoted as τ_d1_, τ_d2_ and τ_e1_, τ_e2_, respectively. The functions P_d_ and P_e_ satisfy the conditions:

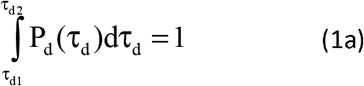

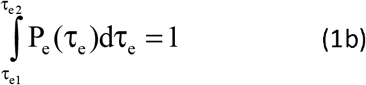

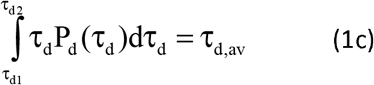

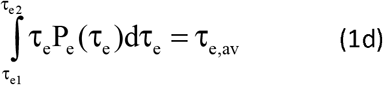

where the subscript “av” denotes the average value of the corresponding variable. Taking into account the above definitions, it can be shown that the functions F(t) and f(t) can be related to each other as:

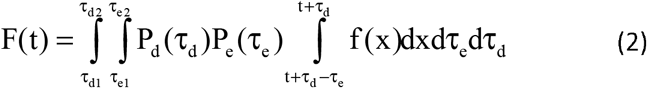

This relation is actually the mathematical expression of the statement that infected persons at time t include those already detected and those to be detected in the next days. In principle, a multiplication of this number by the average shedding rate per person would give the required global shedding rate evolution function R(t). Such averaging over all infected individuals would have been sufficient for data reduction purposes only if the shedding rate of individuals during their disease cycle had a constant value. However, as already explained, this is not true since there is a strong variation in virus concentration in stool not only among infected individuals but also across the days of their disease cycle.

Let us denote this variable function of the daily shedding rate per person as S(τ), (τ: day of the disease onset). It has been shown that not only S is not a constant, but on the contrary, it is a strong function of its argument τ. In order to incorporate the effect of function S(τ) to the global shedding rate R(t), knowledge of the distribution of disease days among the infected population is required. The corresponding function is denoted as F*(τ,t) and represents the density function of the distribution of disease days, τ, at time t. It is fruitful to decompose the function F*(τ,t) into the product of the total number of infected people and the probability density function of the days of infection as F*(τ,t)=F(t)g(t,τ) where g(t,τ) is a probability density function with respect to τ and satisfies the condition:

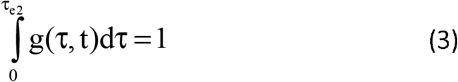

It can be shown that the function g can be derived as

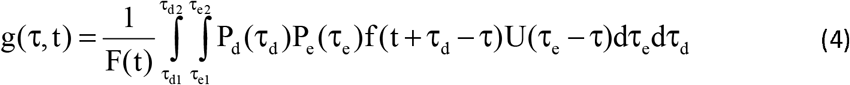

where U is a step function taking the values 0 for negative and 1 for positive argument. The domain of definition of the above expression is from t=τ_e2_-τ_d1_ to t=T-τ_d2_ in the case of f(t) known from t=1 to t=T.

### Parametrization of the function S(τ)

In order to proceed, one should know the function of virus shedding rate in stool per person and per day of the disease. A first question is how the onset of infection (i.e., time τ=1) is defined. There are several possibilities for this definition but in the present context the most relevant one is that τ=1 day is the first day of non-zero shedding of virus in the stool of a person.

There are only a few available clinical studies in literature (e.g. Wölfel et al.(Wölfel et al., 2020), Huang et al.(Huang et al., 2020); Tan et al.(Tan et al., 2020)) referring to the kinetics of SARS-CoV-2 shedding rate in stool during the course of disease (from τ=1 to τ_e_). To our best knowledge, from those studies only the work of Wölfel et al.(Wölfel et al., 2020) reports actual viral concentrations whereas all other studies refer to Ct values from molecular analysis. It must be mentioned that these studies present data exclusively from hospitalized patients, thus from people presenting moderate to severe symptoms. Apparently, it is not easy to collect stool samples from infected persons prior the onset of symptoms, and as a result, asymptomatic or mild cases are not registered in clinical studies. A first common observation in these studies is that virus concentration in stool among infected persons vary by several orders of magnitude. Therefore, here an average shedding function per person will be employed and the significance of variations among individuals will be discussed on statistical terms. Interestingly, many past studies assumed in addition a uniform with respect to time shedding rate despite the orders of magnitude variability across the course of the disease (Ahmed et al., 2020; Gonzalez et al., 2020; Medema et al., 2020a; Saththasivam et al., 2021). This assumption simplifies the algebra of the problem enormously but it is incorrect.

A thorough fitting procedure of the virus titers data in stool of Wölfel et al.(Wölfel et al., 2020), led to a peculiar two-steps model(Miura et al., 2021): a first step with zero viral shedding, where virus load simply accumulates in infected hosts up to a maximum concentration reached at the day of symptoms onset, and a subsequent second step characterized by viral shedding at an exponentially decreasing concentration over the days of the disease. This fitting yields a Gamma function which degenerates to an exponential one. Evidently, it is quite arbitrary to assume that there is an initial viral accumulation period up to a maximum concentration without any shedding at all.

Here, an even more general parameterization is introduced. The average over the infected persons function is represented as a product of the following factors: (1) the average stool amount produced by a person per day, A (g_stool_/day), (2) the maximum in time (averaged over infected individuals) virus concentration in stool, B (g_virus_/g_stool_) and (3) a time distribution function s(τ) of this concentration. This distribution is assumed to consist of an exponential increase from a minimum initial value up to a maximum value being followed by an exponential decrease down to a minimum final value. These minimum initial and final values of the distribution are assumed to be 1% of the maximum value. As a result, the whole distribution spans shedding rates over two orders of magnitude. This assumption may be changed to any other option, e.g., to 1‰ (three orders of magnitude span), but the approach remains the same. However, a two orders of magnitude span represents adequately most of the shedding of infected individuals during a typical course of the disease, so it is adopted herein. A new parameter is introduced, τ_a_, which denotes the value of τ days at which the maximum virus concentration in stool, i.e. maximum shedding rate, occurs. Summarizing, the above the function

S(τ;τ_a_,τ_e_)is given as:

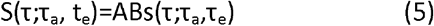

Where s(τ;τ_a_,τ_e_)is

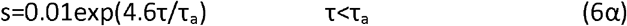

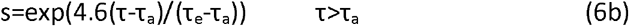

The shape of the function s(τ) for several pairs of (τ_a_,τ_e_) - (6,32), (12,32), (9,25) - is shown in **Figure 1**. The particular parametrization of the function S proposed in the present work is more realistic than the Miura et al. (Miura et al., 2021) model, as it incorporates viral shedding even in the early days of the disease and before the peak maximum (peak) viral concentration in stool is reached. Such early shedding in stool is in line with clinical studies reporting patients with shedding in oropharyngeal swabs (He et al., 2020) and gastrointestinal symptoms (Siegel et al., 2020) a few days before the appearance of respiratory symptoms. Nevertheless, one should keep in mind that respiratory shedding is only a proxy for fecal shedding. In addition, the proposed function S is very attractive because there is one to one correspondence between the parameters and major features of the function. The height (amplitude) of S is determined by the product AB, its time length is determined by the parameter τ_e_ and its skewness by the parameter τ_a_. In particular, the closer τ_a_ is to τ_e_/2 the smaller the skewness is. It must be stressed that the proposed parametrization of S does not require explicit information on the duration of the incubation period after infection nor on the day of symptoms onset. What actually matters in the context of the present analysis is the day of the maximum shedding rate as this is described by the parameter τ_a_. Some clinical studies indicated that this maximum might occurs at the end of the incubation period which is taken also as the day of symptom onset (Huang et al., 2020; Miura et al., 2021; Tan et al., 2020). However, in the present formulation this day is flexible and can be anytime from the moment of infection to the end of the shedding period (i.e., the active days of the disease).

**Figure 1.**
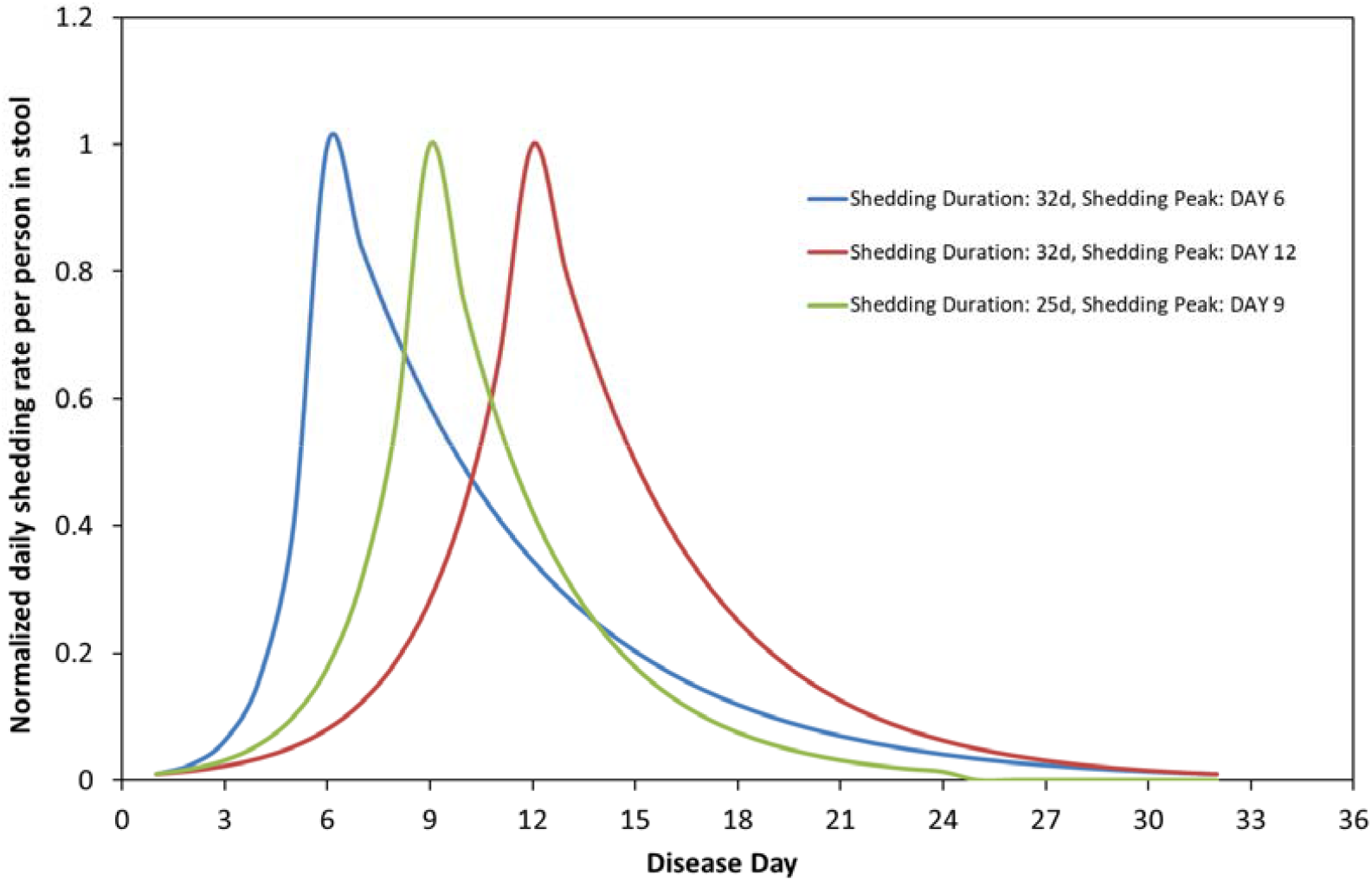
Time distribution of normalized shedding rate per person in stool at different shedding durations and shedding peak days.

The final expression for the global shedding rate is taken by integrating the individual’s shedding rate over the days of the disease.

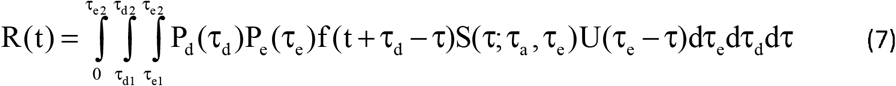

The development up to now is rather complex and includes several unknown probability density functions. In the absence of any information about them, it is convenient to consider them as Dirac delta functions. In this way, we assign to each distribution its average value. This approach not only simplifies considerably the mathematical problem but it also allows -through sensitivity analysis- to assess bounds on the effect of using a different distribution than the Dirac delta function. This is based on the principle that any secondary feature of a distribution has much smaller effect on the result than its average value. By considering the relations P_d_(τ_d_)=δ(τ_d_-τ_d,av_) and P_e_(τ_e_)=δ(τ_e_-τ_e,av_) (where δ denotes the Dirac delta function) and substituting them in equations (2), (4), (7) leads after some algebra to (the subscript “av” is dropped in the following for clarity):

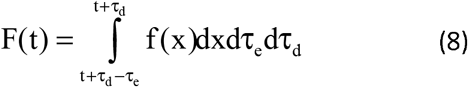

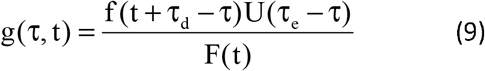

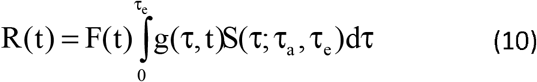

The above set of equations is discretized to be compatible with the present data. These data refer to daily values of virus shedding rate so a finite volume discretization is followed here. Let us denote as f_i_ the number of positive medical tests at day i (measurement period i=1 to N), F_i_ is the total number of infected people (that is daily cases) at day i (with the specimen of their positive test collected at day i), R_i_ the shedding rate at day i and g_i,j_ the probability of being at the j-th day of the disease at the calendar day i. The governing equations take the form:

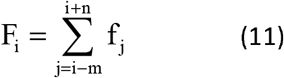

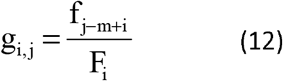

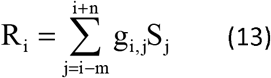

for i>m and i<N-n

where S_j_ is the shedding rate per person at the j-th day of the disease (m=τ_e_-τ_d_, n=τ_d_)

## RESULTS

Figure 2. displays medical surveillance data reported for the city of Thessaloniki (∼ 700,000 inhabitants) (National Public Health Organization, 2020). More specifically, it presents the daily number of infected people versus the date of their specimen collection for medical testing. The date of specimen collection is back-dated by 1 to 4 days (median of 3 days) from the date of reporting by the Hellenic National Public Health Organization. The difference between the two characteristic dates is small but the epidemiological data adjusted to the date of specimen collection are more appropriate to compare with wastewater measurements because they are better associated with the date of infection and the date of symptoms onset. Nevertheless, if early warning by wastewater measurements is the stake then comparisons should be made with medical surveillance data as announced, i.e., based on the date of reporting.

**Figure 2.**
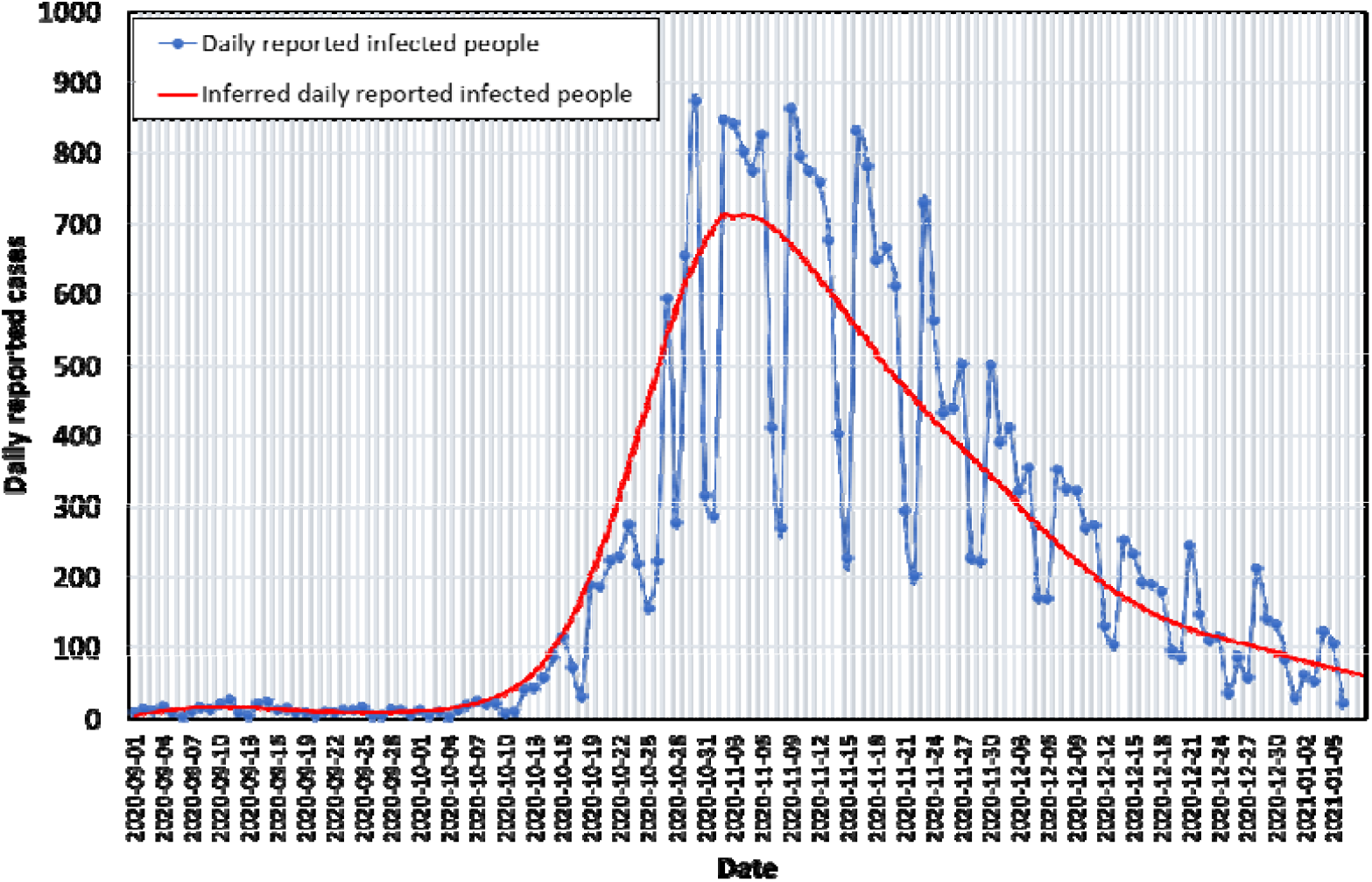
Medical surveillance data reported for the city of Thessaloniki from September 1^st^, 2020 to January 6^th^, 2021 and corresponding Bayesian model fit curve.

The presented data cover the period from September, 1^st^ 2020 to January 6^th^, 2021. A fitting curve is fitted over the raw data to smooth out the noise. Most of the parametric study that follows is performed with respect to this curve, to skip the noise. Following the loose atmosphere of Summer 2020, in September and October there was no strict quarantine in the city and only a modest rule for social distancing was active along with a rule for limited number of people in confined places, like stores and restaurants. In September and the first 10 days of October the daily reported cases were always well below 20 and often even below 10. At around mid-October 2020 the number of infected cases started to escalate exponentially. In just two weeks, the number of cases increased about ten-fold in just two weeks. This dramatic rise was consistent with the about 2.7 days doubling time of the epidemic reported for European countries in the absence of control measures (SET-C Steering Committee, 2020). A strict lock-down was imposed on the city on November 2^nd^ meant to suppress the outburst of the disease. Between Nov 3^rd^ and Nov 17^th^ the number of infected cases fluctuated approximately from 800 to 300 cases between weekdays and weekends. Similar fluctuations were noticed also in other studies (Peccia et al., 2020). On December 12^th^ some of the strict measures were released, e.g., retail stores opened to serve people but only at their entrance door and only through pre-scheduled appointments. As a result, after mid-November the number of infected cases gradually went down until it became pretty stable at below 100 daily cases in the first week of 2021.

Figure 3. presents the experimentally determined relative shedding rate of viral RNA copies, r_1_(t)=R_exp_/R_expo_, in sewage from October 5^th^, 2020 to January 6^th^, 2021 for Thessaloniki. October 5^th^ is the last day after summer that the measured viral RNA copies in wastewater fluctuated around the experimental limit of quantification of the employed technique (∼10 viral RNA copies/mL). R_exp_ represents the daily value of shedding rate, whereas R_expo_ is a reference shedding rate. Presenting experimental shedding data in the form of a ratio reduces the effect of determination uncertainty. R_expo_ has been defined as the average R_exp_ value across the days of the first week of October which was a period with calm epidemiological conditions in the city (less than 10 daily reported cases).

**Figure 3.**
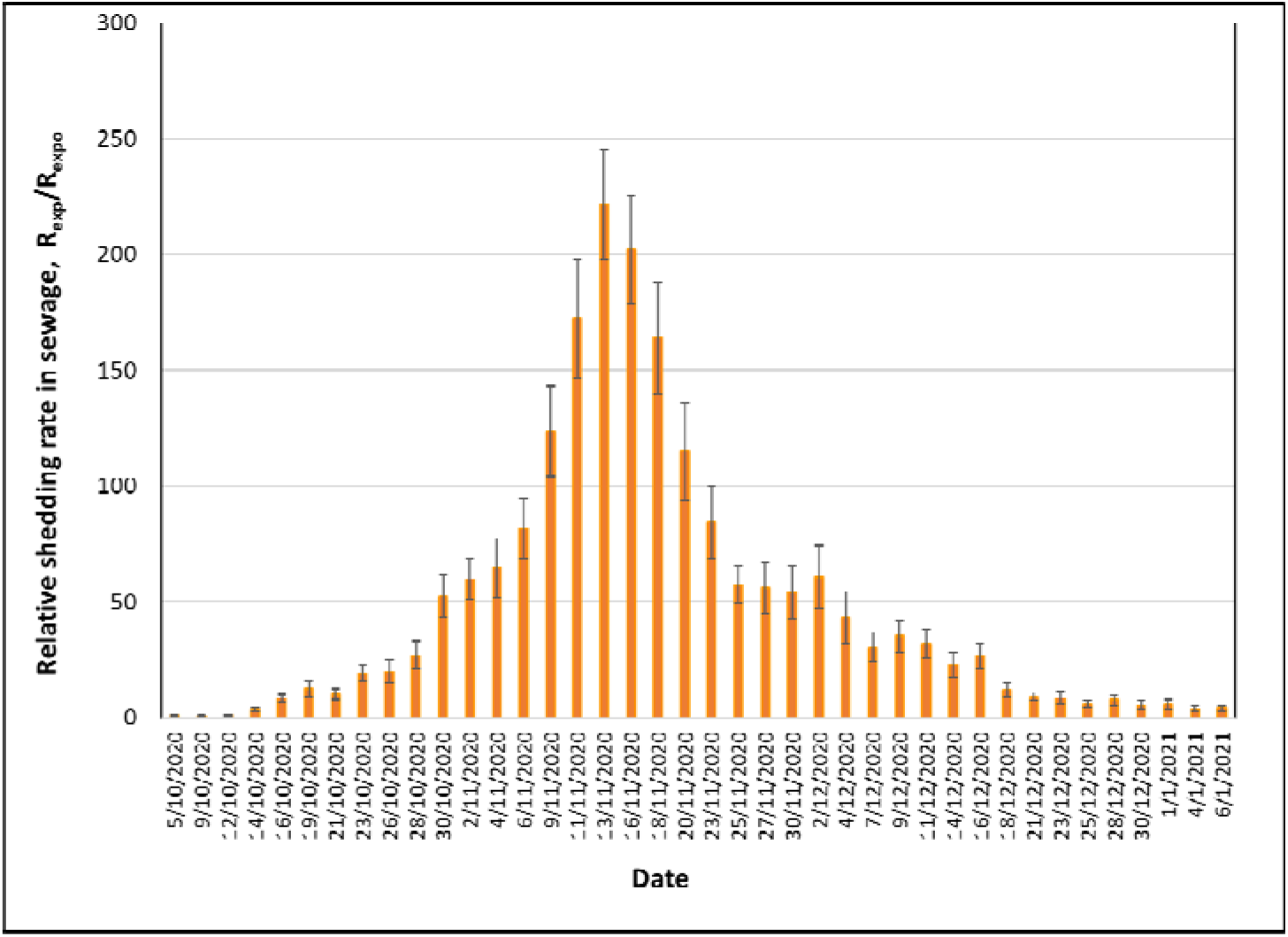
**E**xperimental relative shedding rate in sewage expressed as the ratio of the measured daily shedding rate (R_exp_) over the reference shedding rate (R_expo_) from October 5^th^, 2020 to January 6^th^, 2021.

There are certain similarities and differences between the medical surveillance data in Figure 2 and the wastewater data in Figure 3. They both show an initial abrupt ascend followed by a later gradual decline. They both show a peak in the first half of November 2020. Yet, the peak in wastewater data is sharp at around November 13^th^ whereas the peak in medical surveillance, despite the intense noise, looks like a plateau between November 3^rd^ and 12^th^. The initial increasing slope in October’s data in these Figures is a bit steeper for wastewater data implying that wastewater surveillance may act as an early warning indicator for medical surveillance. More about this below.

Next, the time series of the reported daily infected cases in the city of Thessaloniki is analyzed using the procedure in the previous section to determine the functions F, g, R. It is reminded that these three functions correspond respectively to (i) the total number of infected people with positive test at a calendar day, t, (those already registered as positive to day t but also those that will be tested in the next days and their test result will be also registered to day t), (ii) the distribution of the number of infected people with respect to the days of the disease, τ, at a calendar day, t, and (iii) the total shedding rate with respect to the calendar day, t. It is important to note that since the temporal discretization quantity is one day, the continuous and the discrete forms of the above functions are arithmetically similar. Thus, the presented results can be read both as continuous functions (per day) or as discrete values (i.e., histograms). The essence of the proposed approach is that it accounts for the time dependence of the shedding rate during the disease. Yet, this is important only when the distribution of the daily shedding rates is not uniform among the population, but it evolves along the days of the disease.

Before a parametric analysis is performed it is useful to identify realistic range of values of the model parameters in order to examine their influence. It is reminded that the three model parameters are (i) the total number of shedding days counted from the infection day, τ_e_, (ii) the day of the maximum shedding rate, τ_a_, and (iii) the day of detection (specimen collection), τ_d_. The analysis of the data of Wölfel et al.(Wölfel et al., 2020) by Miura et al(Miura et al., 2021) indicated an average shedding period of 26 days after the onset of symptoms. This is a bit longer than the 17 and 18 days, reported by Huang et al.(Huang et al., 2020) and Tan et al.(Tan et al., 2020), respectively. But both the latter studies reported a high variance in their average shedding period plus they mentioned that in some individuals shedding exceeded five weeks. Apart from this, Tan et al.(Tan et al., 2020), indicated a 6 day median incubation period from the day of infection until the day of symptoms onset. If one considers that the day of symptoms onset is a good candidate for the average day of maximum shedding rate then τ_a_= 6. For Greek patients it is realistic to assume that specimen collection for medical testing takes place on the average about 3 days after the onset of symptoms. This is in line with information in literature that the median time period from symptom onset to hospital admission is 3 days(Huang et al., 2020). Therefore, a realistic average day of detection is τ_d_=8. The above information combined implies a total number of shedding days τ_e_=32 (6+26) and this is the value adopted herein. Summarizing, the base case parameter values around which parametric analysis is performed are τ_a_= 6, τ_d_=8 and τ_e_=32.

Figure 4. shows the computed values of g(τ,t) for selected values of calendar days, namely, September 25^th^, October 10^th^, October 23^th^, December 9^th^ and December 29^th^, 2020. The small oscillation appeared is due to the oscillations in the curve f(t). However, as f(t) – the positive tests count density-increases new patients are added to the number of infected people. The addition occurs at a higher rate than the rate of withdrawal of cured people. This leads to the accumulation of infected people at early shedding days, τ, so the function g(τ) acquires the characteristic, decreasing with τ, profiles shown for October 10^th^ and October 23^th^. In the time regime of the maximum f(t), e.g. curve for December 9^th^, the function g(τ) tends fast towards uniformity, like at the early shedding days. Finally, during the decreasing period of f(t), e.g. curve for December 29^th^, the withdrawal of cured people rate overwhelms the entry of new patients so g(t) is an almost linearly increasing function of τ. The above description manifests that the function g(τ) varies a lot with calendar time t and it is far from uniform along the shedding days so the present analysis is necessary for the estimation of shedding rate across calendar days. It is clear that at a particular calendar day, infected people are at a different stage of the disease, and thus, at a different stage of viral shedding.

**Figure 4.**
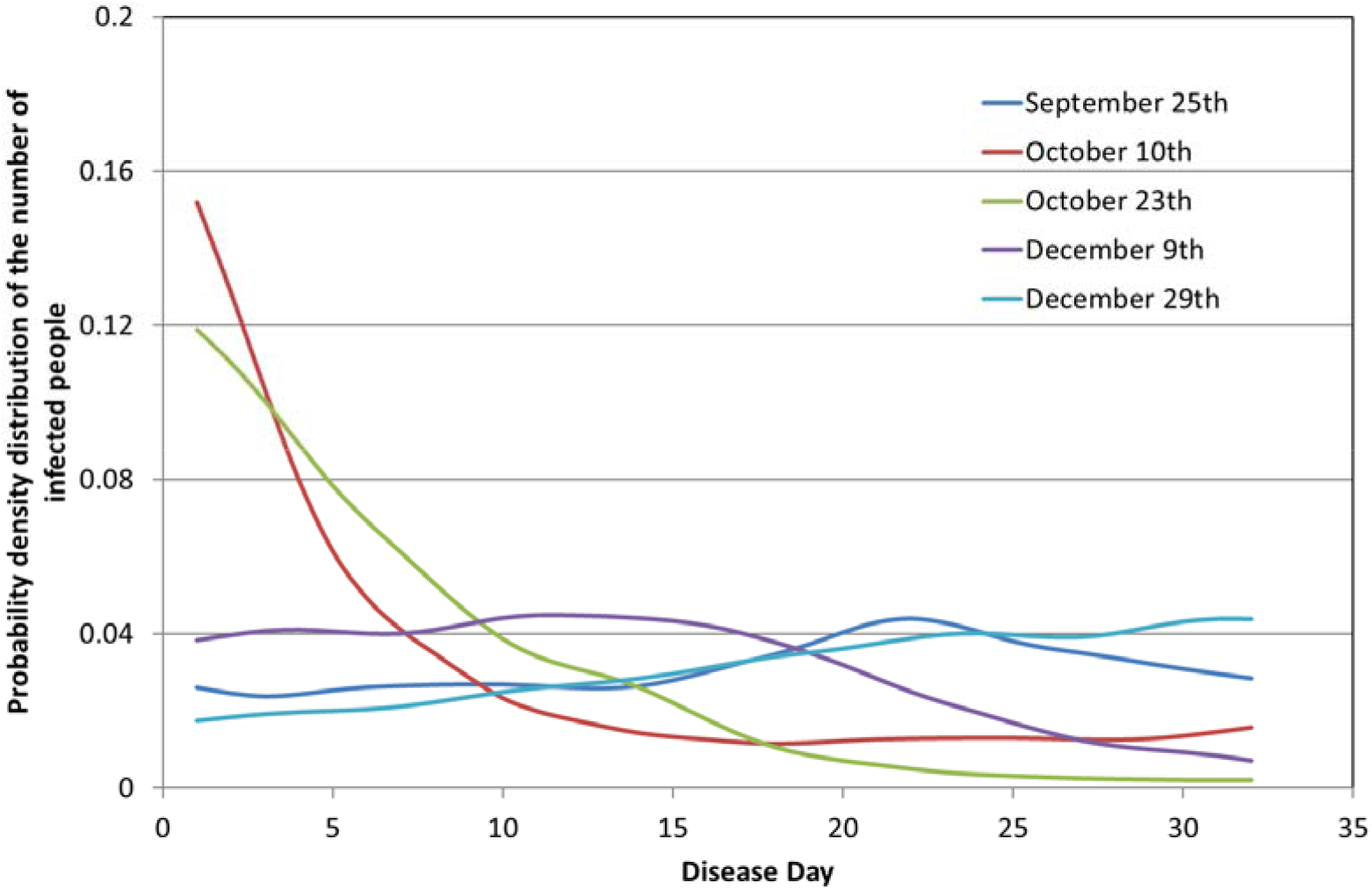
The probability density distribution of the number of infected people with respect to the days of the disease.

The early warning capacity of wastewater surveillance with respect to medical surveillance is estimated by comparing in **Figure 5** the dynamics of functions f(t): daily reported infected people, F(t): total reported infected people, and R(t): experimentally determined viral shedding rate in sewage. More specifically, **Figure 5a** shows this comparison for the base case τ_a_= 6, τ_d_=8, τ_e_=32; **Figure 5b** for τ_a_= 2, τ_d_ =8, τ_e_ =32; **Figure 5c** for τ_a_ = 12, τ_d_ =8, τ_e_ =32 (all values in days). Curves in **Figure 5** stop at December 29^th^, 2020, and not at January 6^th^, 2021. This is because calculations of F(t) -and accordingly of R(t)-can be performed only up to December 29^th^, 2020, as this function requires for every daily value available data for the subsequent τ_d_=8 days. To avoid experimental noise in the parametric analysis, calculations in **Figure 5** are based on the Bayesian fitted curve in **Figure 2**. For the comparison it is imperative to express these three functions in a similar scale. This is done by transforming them in their probability density function counterparts. For the present case, this is done simply by dividing each function with the sum of its values for the period of interest. The function F(t) always resides after f(t) which is typical for a cumulative type of function. The temporal distance apart between these two functions depends on the values of the parameters τ_d_ and τ_e_. This distance increases as τ_d_ decreases and τ_e_ increases. The dynamics of the function R(t) depend also on τ_a_. In case of τ_a_<τ_d_ then R(t) precedes f(t). This condition is extremely important as it demonstrates that for wastewater surveillance to precede medical surveillance the day of the peak of viral shedding rate in stool during the disease days must lie before the day of specimen collection for medical testing. In all other cases, the curve R(t) resides between f(t) and F(t), with the latter being the approximate long time bound for the dynamics of R(t). The previous statement can be confirmed by observing the three aforementioned functions for the three set of parameters in **Figure 5**. Depending on the calendar day, function R(t) precedes f(t) roughly from 0 to 4 (b) days **(Figure 5b)**. For the base case in **Figure 5a** the difference between R(t) and f(t) appears marginal. Yet, one must recall that in **Figure 2** the ascending part of the Bayesian curve precedes raw data, so in reality even for the base case sewage data lie earlier than medical data. In literature there is experimental evidence of identifying SARS-CoV-2 in wastewater earlier than medical reporting by several days, e.g., 2 days in Ottawa Canada(D’Aoust et al., 2021), 2-4 days in Montana USA(Nemudryi et al., 2020). In summary, the viral shedding rate evolution curve measured in wastewater lies from a few days before the curve of the number of daily infected people up to the curve of the total number of infected people. The exact position of the curves depends on the relation between the day of maximum shedding rate in stool and the day of specimen collection for medical testing.

**Figure 5.**
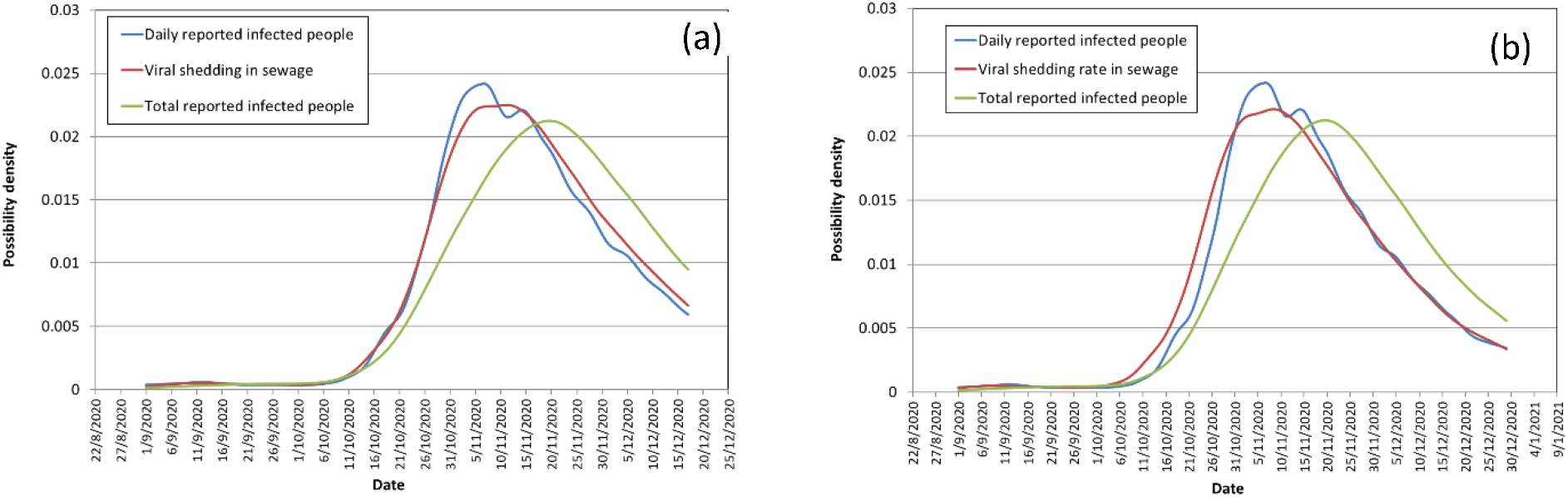
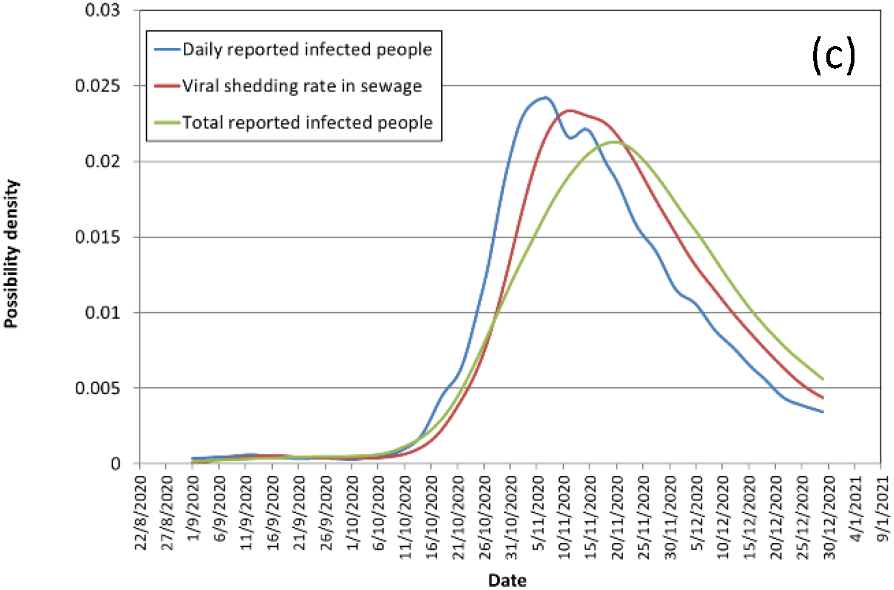
Comparison of the relative viral shedding rate in sewage with the number of the daily and total reported infected people per surveillance day. In all cases shedding duration in stool is set as 32days, laboratory test is assumed at DAY 8, whereas the day of shedding peak in stool is at: DAY 6 (a), DAY 2 (b) and DAY 12 (c).

For appraising in real life the early diagnostic capacity of wastewater surveillance it is important to compare wastewater data with medical surveillance data based on the date of reporting (and not on the date of specimen collection). Peccia et al.(Peccia et al., 2020) found in New Haven (USA) that although wastewater data was ahead by only 0-2 days of positive test results by the date of specimen collection, it was 6-8 days ahead of positive test results by the reporting date. In Thessaloniki (National Public Health Organization, 2020) the date of reporting follows the date of specimen collection by about 2 days on the average and so even the base case parameters in **Figure 5a** permit an early diagnosis capacity of wastewater surveillance of around 2 days.

A critical issue in the disease dynamics is the effect of the number of unreported infected people (unreported cases). Let us denote as U the ratio of unreported to reported infected people. If U is constant along the days of the shedding period, the analysis is exactly the same with that shown above for the reported infected people, F, since the probability density functions do not change. However, if the number of unreported people varies along the shedding period, then this may yield a time lag between wastewater and medical data. **Figure 6** compares the relative medical surveillance data of the smoothed (Bayesian fit) daily reported cases, f(t)/f_o_, versus the theoretically estimated relative shedding rate in wastewater, r_2_(t)=R(t)/R_o_. Normalization parameters, f_o_ and R_o_ are average values of the respective parameters over the same reference first week of October. Comparisons are for the base case τ_a_= 6, τ_d_=8, τ_e_=32. The ratio U is assumed to vary proportionally with the number of reported infected people. The parameter U_max_ is used to parameterize U designating the maximum value of U for the case considered. The parameter U_max_ takes the value 0.5, 2 and 4 in **Figure 6**. Apparently, for U_max_=0.5 (unreported cases becomes at most 50% of reported cases) wastewater and medical data coincide at the ascent of the curves but at the descent wastewater lags behind. This changes for U_max_=2 (unreported cases becomes at most 200% of reported cases) and for U_max_=4 (unreported cases becomes at most 400% of reported cases) where wastewater data clearly go before medical data at the ascent of the curve. Summing up, a rising number of unreported cases as the shedding rate increases leads to earlier wastewater signal than medical surveillance. Furthermore, the higher the maximum number of unreported cases the more in advance the wastewater data from the medical data.

**Figure 6.**
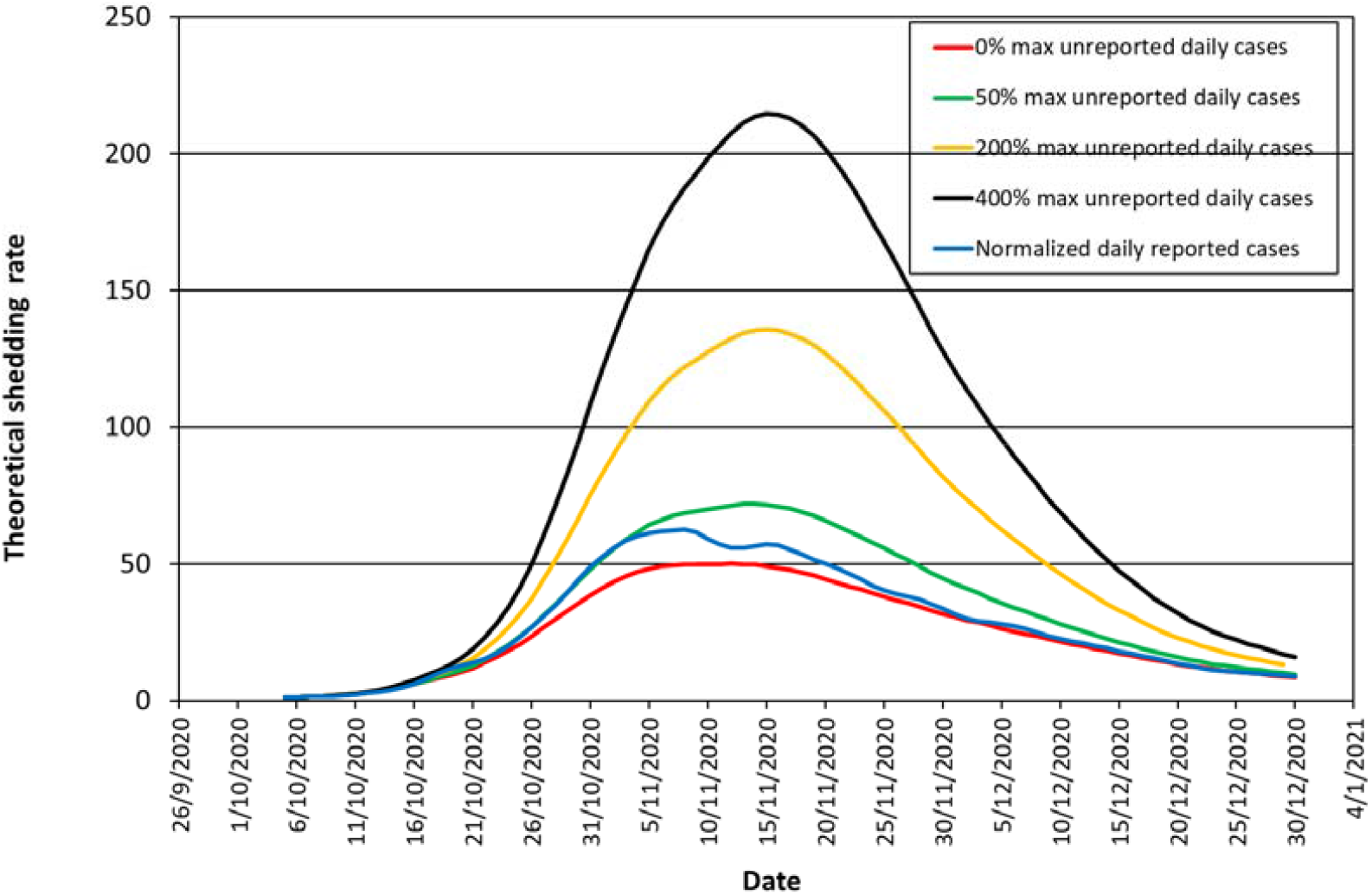
Comparison of the theoretical relative shedding rate in sewage with the relative medical surveillance data of the smoothed (Bayesian fit) daily reported cases for different scenarios of unreported daily cases. Unreported daily cases vary proportionally with the number of reported daily cases starting from 0 and reaching a maximum value of 0, 50, 200, 400% of reported cases at the date of maximum shedding rate.

Next, the experimental relative shedding rate, r_1_(t)=R_exp_(t)/R_expo_, shown in **Figure 3** is compared with the estimated total number of infected people, including both reported and unreported ones, **Figure 7**. Red circles denote increasing shedding rates whereas green squares decreasing shedding rates, respectively. It is apparent that from low to moderate relative shedding rates (R_exp_(t)/R_expo_ <100) and total number of infected people (F<15000) there is a roughly linear relationship between the two quantities. This changes dramatically for higher values of either quantities. Moreover, during the increasing phase of the relative shedding rate, i.e., outbreak of pandemic wave, the same number of infected people sheds a higher viral load than during the decreasing phase. This is so because at the outbreak phase of the pandemic most infected people are at the early phase of their disease when shedding rate is higher, as shown in **Figure 1**. In line with this, when the relative shedding rate starts to fall right after its peak, the total number of infected people continues to rise to about 10% more cases. This happens because the turning point (peak) of the curve designates the moment when the daily new infected cases begin to reduce, **Figure 2**, so the majority of the total number of infected people are at later phase of their disease when shedding rate is lower.

**Figure 7.**
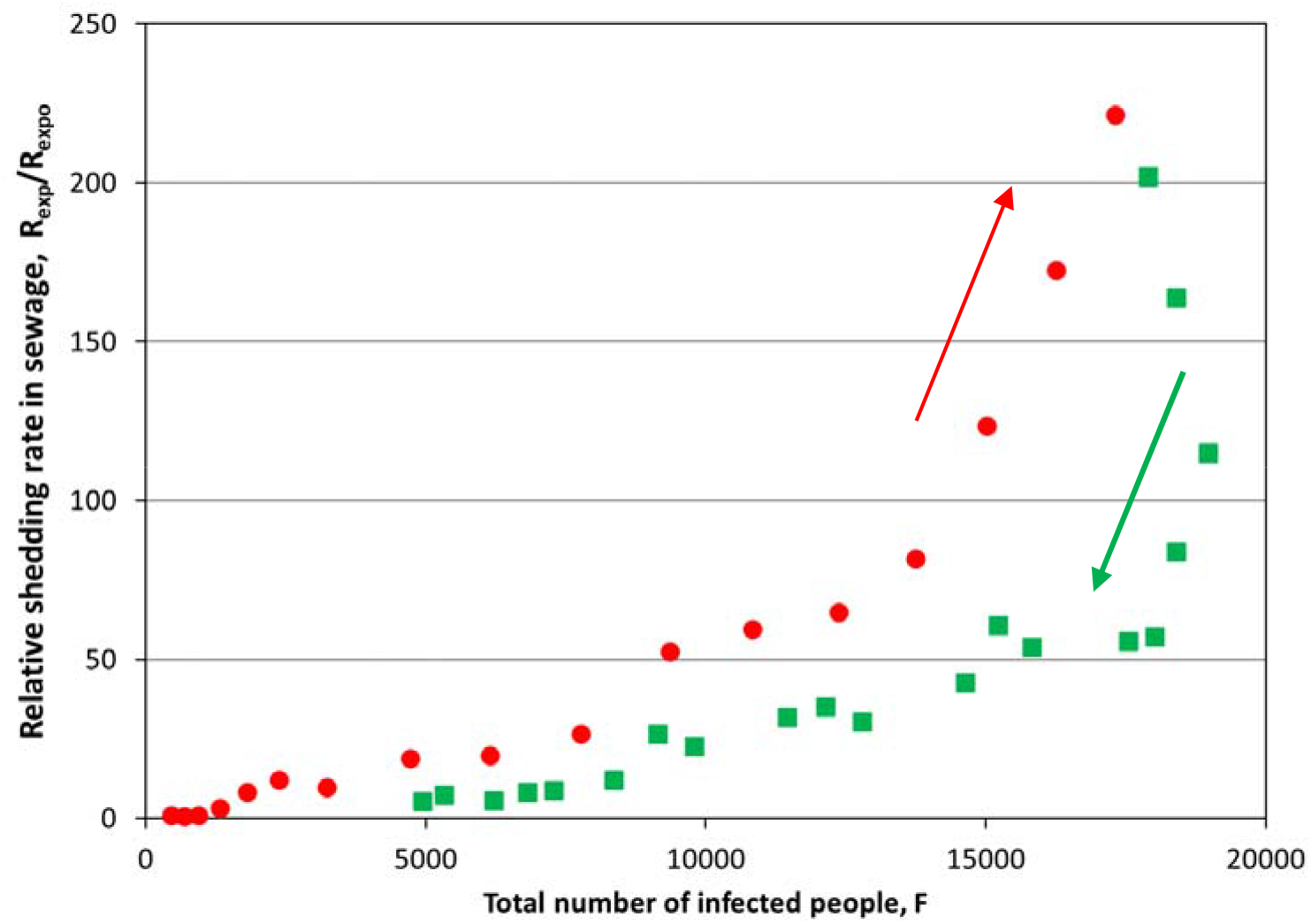
Experimental relative shedding rate in sewage, R_exp_(t)/R_expo_, versus the estimated total number of infected people (reported and unreported), F. The latter is estimated assuming that, on the average, the shedding duration in stool is 32 days and medical tests are taken at DAY 8 during the disease. Red circles denote increasing shedding rates whereas green squares denote decreasing shedding rates.

Let us now try to correlate the theoretically derived function R(t) with the actual one found from experimental wastewater analysis R_exp_(t). The latter was shown as a relative shedding rate, r_1_(t)=R_exp_(t)/R_expo_, in **Figs 3** and **7**. Contrary to what was done in **Figure 6**, now the theoretically estimated relative shedding rate r_2_(t)=R(t)/R_o_ is computed from the noisy medical surveillance raw data in **Figure 2** for the base case parameter values τ_a_= 6, τ_d_= 8 and τ_e_=32. As a result, r_2_(t) data are also noisy. Therefore, β(t)=r_1_(t)/r_2_(t) is a ratio reflecting the total number of reported and unreported infected people, F(1+U), over the total number of reported infected people, F, at every calendar day. In other words, β=1+U, that is, β-1 stands for U, the ratio of the total unreported over the total reported cases. Thus, the minimum value of β is 1 and corresponds to zero unreported cases. As before, the values of R_expo_ and R_o_ both refer to the same reference first week of October. Selection of the reference week at an epidemiologically calm period allows assuming that R_expo_/R_o_≈1 so the normalization in β can be ignored. The evolution of β with the calendar days appears in **Figure 8**. Considering the intense day-by-da scatter, the ratio β takes values roughly between 1 and 5, following a qualitatively similar trend with the wastewater shedding data in **Figure 3**. Interestingly, these results imply that there are essentially no unreported cases at periods of low shedding but unreported cases rise up to four times the reported ones (U/F≈4) when shedding is maximum. Analysis of seroprevalence data in Qatar indicated that diagnosed infections represented about 10% of actual cases(Saththasivam et al., 2021). Another serological-testing study(Havers et al., 2020) showed that the actual number of infections could be from 6 to 24 times the number of the reported cases. Overall, the presently estimated values of U/F from ∼0 to 4 is close to the above published values. To our knowledge, this is the first time in the SARS-CoV-2 pandemic that wastewater data are employed to indicate a proportion between unreported and reported infected cases. Moreover, the present study shows that the proportion of unreported to reported cases is not constant during the outbreak of a disease wave but as the number of infected people (and shedding rate) increases, it attains higher values and its scatter decreases.

**Figure 8.**
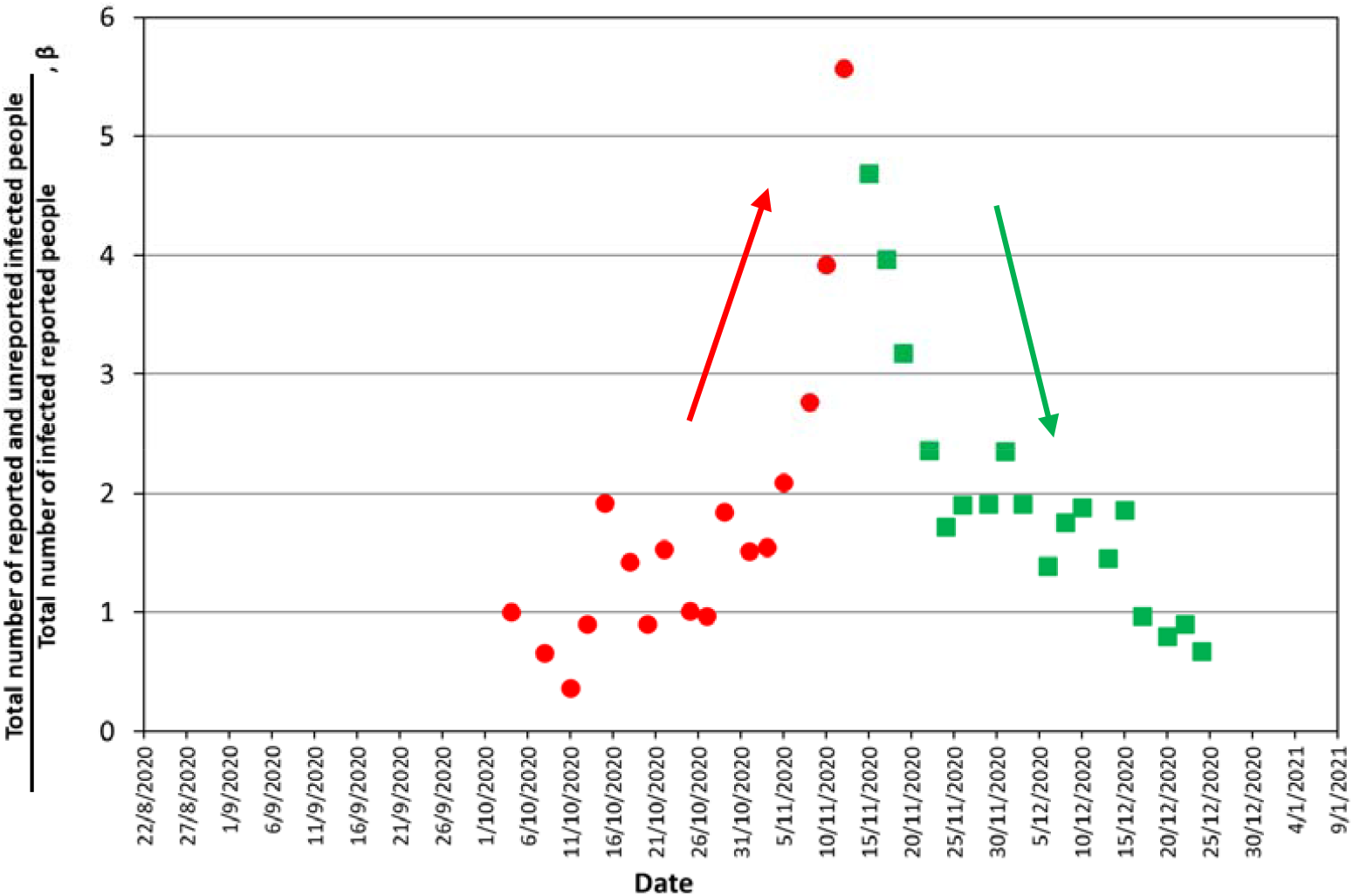
Evolution of the ratio of the total number of infected people (reported + unreported) over the number of reported infected people. The ratio is calculated from unsmoothed raw data in Figures 2 and 3. Red circles denote increasing shedding rates whereas green squares denote decreasing shedding rates.

Based on the above, an effort is made to correlate β to the total number of the reported infected cases F. The result is shown in **Figure 9a** where once more red circles denote increasing shedding rates and green squares denote decreasing shedding rates. Alike in **Figure 7**, β starts decreasing before the maximum F is reached. In addition, at high F values there is a hysteresis in β between increasing and decreasing branches of the curves. This might imply that virus spread among unreported people is a bit higher in the declining phase of the disease either because of the cumulatively higher virus prevalence in the community or because more people undergo testing during the ascending spreading period of the disease than in the milder descending period. **Figure 9b** presents β versus the daily reported number of the infected cases, f. Interestingly there is an almost linear relation between the two parameters up to about 400 daily reported cases. After that point, spreading of the virus in the community is too high so β depends on the total number infected people F and not on the daily reported cases.

**Figure 9.**
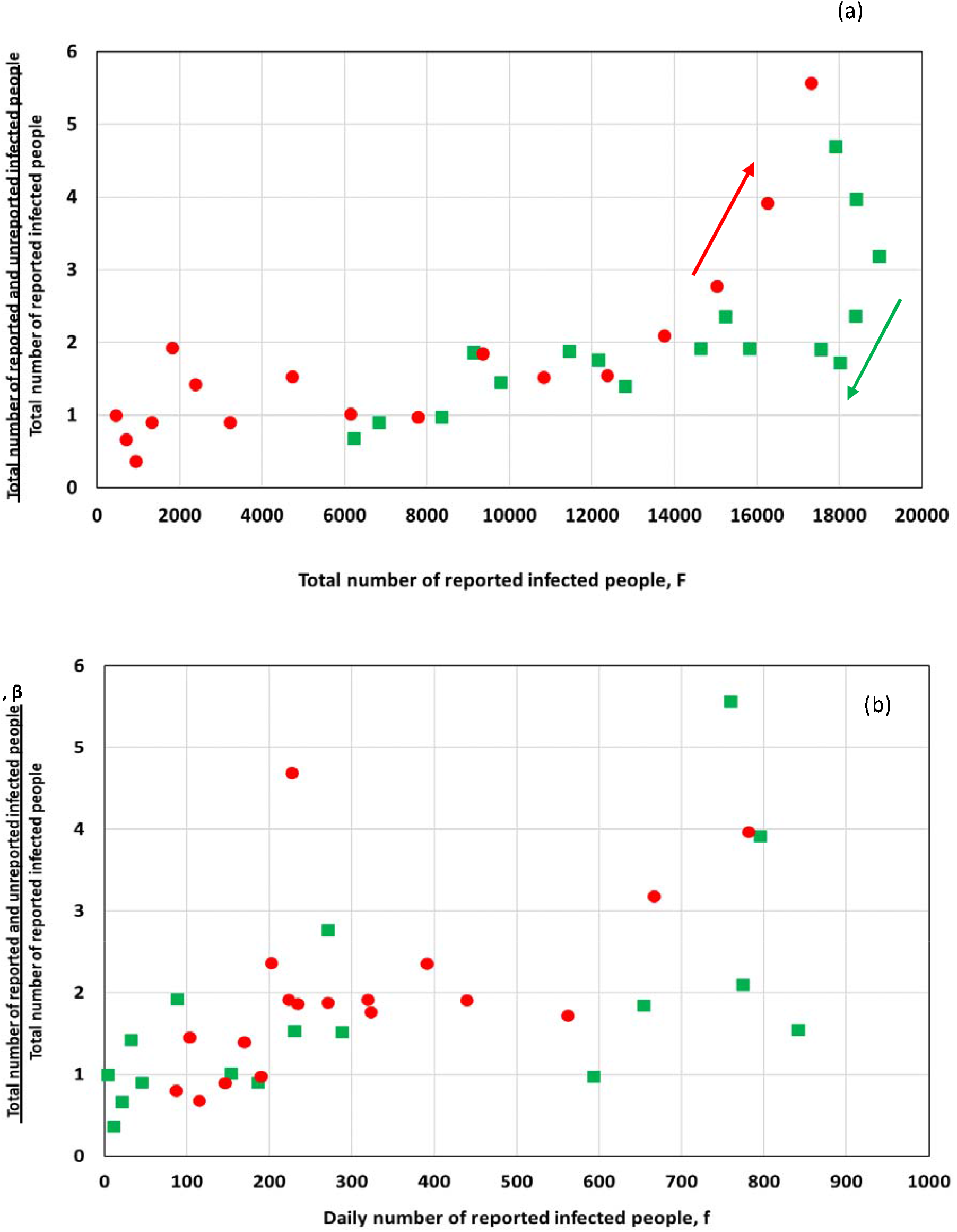
Variation of the ratio of the total number of infected people (reported + unreported) over the number of reported infected people with regards to (a) the total number of reported infected people and (b) the daily number of infected people. The ratio is calculated from unsmoothed raw data in Figures 2 and 3. Red circles denote increasing shedding rates whereas green squares denote decreasing shedding rates.

It must be mentioned that β may depend also on other quantities (apart from F and the ascending/descending mode), like the value of the F slope, but at present there are not enough data to support this. Having an estimation of the total number of infected people through β, a crude estimation of the product AB (average maximum virus shedding rate per person) is obtained. The estimated value of AB is at about 2 ·10^11^ day^-1^. This value is in fair proximity with that estimated by the wastewater analysis of Wu et al.(Wu et al., 2020). However, more work is needed to allow judging on the correctness of this value.

## CONCLUSIONS

A mathematical model is developed that permits estimation of SARS-CoV-2 shedding rate in wastewater from the number of daily infected people (daily cases) announced by medical surveillance. The problem is complex as it requires a cumulative function of the total number of infected people at every calendar day and a function of the average shedding rate among infected individuals at every day along the course of the disease. Based on the limited available evidence in literature, a realistic function for the shedding rate of SARS-CoV-2 in stool of infected individuals is proposed which calls for an exponential increase of shedding rate from the day of infection to the day of symptoms onset, being followed by an exponential decay until the end of the disease. Three characteristic times describe this function: the day of maximum shedding rate, the day of detection (specimen collection) and the end day after the onset of the disease. Applying this model to the public health surveillance data for the city of Thessaloniki (∼700,000 inhabitants, North Greece) a thorough parametric study is performed. It is shown that for WBE to afford an early warning capacity, the day of maximum shedding rate must precede the day of detection by a few days. In particular, at the beginning of an outbreak curve, a 4-day early WBE signal requires these two characteristic times to be apart by 6 days. For Thessaloniki where on the average these two characteristic times are apart by just 2 days but, additionally, the day of reporting follows the date of specimen collection also by about 2 days, the early diagnosis capacity of wastewater surveillance is around 2 days. Furthermore, comparison of wastewater surveillance and public health data indicates the existence of a number of unreported infected people. For Thessaloniki in the outbreak of November 2020, this number was negligible at epidemiologically calm days with low shedding rate but went up to four times the number of reported people when shedding rate reached a climax at mid-November 2020. Interestingly, the presence of an increasing number of unreported cases with shedding rate enhances the early warning capacity of WBE. To this end, the present model is an essential tool to investigate the dynamics of virus spreading based on wastewater measurements. When such knowledge is adequately acquired then the inverse problem of estimating the number of cases from wastewater data can be attempted.

## Data Availability

Raw data of wastewater quality characteristics and reposrted cases are provided in supplementary materials

