## Supplementary materials for "Relating SARS-CoV-2 shedding rate in wastewater to daily positive tests data: A consistent model based approach" (from another version of this preprint: version 1, 2021-07-06)

**Maria Petala[1], Margaritis Kostoglou[2], Thodoris Karapantsios[2], Chrystsomos Dovas[3], Theodoros Lytras[4,5], Dimitrios Paraskevis[6], Emmanouel Roilides[7], Anastasia Koutsolioutsou-Benaki[8], , Georgios Panagiotakopoulos[4], Vana Sypsa[6], Symeon Metallidis[9], Anna Papa[10], Efstratios Stylianidis[11], Agis Papadopoulos[12], Sotirios Tsiodras[6], Nikolaos Papaioannou[3]**

[1]Laboratory of Environmental Engineering & Planning, Department. of Civil Engineering, Aristotle University of Thessaloniki, Thessaloniki, 54 124, Greece
[2]Laboratory of Chemical and Environmental Technology, Dept. of Chemistry, Aristotle University of Thessaloniki, 54 124 Thessaloniki, 54124, Greece
[3]Faculty of Veterinary Medicine, School of Health Sciences, Aristotle University of Thessaloniki, Thessaloniki, 54124, Greece
[4]National Public Health Organization, Athens, Greece
[5]European University Cyprus, Nicosia, Cyprus
[6]National and Kapodistrian University of Athens, Athens, Greece
[7]Infectious Diseases Unit and 3rd Department of Pediatrics, Aristotle University School of Health Sciences, Hippokration Hospital, Thessaloniki, 54642, Greece
[8]Department of Environmental Health, Directory of Epidemiology and Prevention of Non Communicable Diseases and Injuries, National Public Health Organization, Athens, Greece
[9]Department of Haematology, First Department of Internal Medicine, Faculty of Medicine, AHEPA General Hospital, Aristotle University of Thessaloniki, Thessaloniki, 54636, Greece
[10]Department of Microbiology, Medical School, Aristotle University of Thessaloniki, Thessaloniki, 54124 Greece
[11]School of Spatial Planning and Development, Faculty of Engineering, Aristotle University of Thessaloniki, Thessaloniki, 54124, Greece
[12]EYATH S.A., Thessaloniki Water Supply and Sewerage Company S.A., Thessaloniki, 54636, Greece

**Table 1.** Physical-chemical characteristics of wastewater samples

| **Date** | **Unit** | **2/10/20** | **5/10/20** | **9/10/20** | **12/10/20** | **14/10/20** | **16/10/20** | **19/10/20** | **21/10/20** | **23/10/20** |
|---|---|---|---|---|---|---|---|---|---|---|
| Flow | m3/d | 143,850 | 140,900 | 141,650 | 147,850 | 157,750 | 158,000 | 147,450 | 142,000 | 139,800 |
| SS | mg/L | 400 | 679 | 637 | 616 | 892 | 835 | 1,083 | 1,084 | 1,019 |
| COD | mg/L | 582 | 1,042 | 1,278 | 938 | 1,284 | 1,144 | 1,509 | 1,480 | 1,518 |
| BOD5 | mg/L | 255 | 429 | 515 | 383 | 492 | 446 | 574 | 583 | 564 |
| NH4-N (Δ) | mg/L | 33.5 | 32.7 | 29.6 | 27.8 | 29.3 | 24.5 | 27.8 | 31.5 | 36.3 |
| NO3-N (Δ) | mg/L | 0.92 | 0.92 | 0.99 | 1.36 | 2.33 | 0.92 | 0.66 | 0.87 | 0.23 |
| TN | mg/L | 64 | 64.4 | 61.5 | 63.9 | 93.7 | 59.4 | 76.2 | 71.8 | 89.4 |
| TOTAL-P | mg/L | | | | 8.99 | 8.92 | | | 13.10 | |
| Conductivity | mS/cm | 7.42 | 7.77 | 8.26 | 7.99 | 4.67 | 3.83 | 7.25 | 6.73 | 5.69 |
| pH | - | 7.74 | 7.56 | 7.63 | 7.57 | 7.59 | 7.52 | 7.60 | 7.48 | 7.65 |
| DO | mg/L | 0.80 | 1.10 | 0.85 | 1.10 | 1.20 | 0.90 | 1.10 | 0.90 | 1.00 |
| UV254 nm | cm-1 | 0.35 | 0.53 | 0.53 | 0.54 | 0.42 | 0.59 | 0.48 | 0.60 | 0.56 |
| DOC | mg/L | 40.00 | 48.00 | 50.76 | 55.50 | 50.00 | 53.14 | 37.20 | 38.90 | 47.20 |

| Date | Unit | 26/10/20 | 28/10/20 | 30/10/20 | 2/11/20 | 4/11/20 | 6/11/20 | 9/11/20 | 11/11/20 | 13/11/20 |
|---|---|---|---|---|---|---|---|---|---|---|
| Flow | m3/d | 127,350 | 142,400 | 136,350 | 112,200 | 155,500 | 130,500 | 131,950 | 133,600 | 134,400 |
| SS | mg/L | 1,180 | 1,009 | 1,094 | 1,139 | 1,149 | 1,362 | 1,252 | 1,038 | 1,287 |
| COD | mg/L | 1,593 | 1,651 | 1,438 | 1,667 | 1,750 | 1,788 | 1,870 | 1,600 | 1,520 |
| BOD5 | mg/L | 656 | 629 | 521 | 573 | 560 | 824 | 689 | 692 | 690 |
| NH4-N (Δ) | mg/L | 32.8 | 30.0 | 31.4 | 29.8 | 29 | 29.8 | 29 | 30.7 | 30.8 |
| NO3-N (Δ) | mg/L | 0.65 | 0.92 | 0.77 | 0.93 | 1.6 | 0.9 | 0.75 | 1.38 | 0.66 |
| TN | mg/L | 87.3 | 74.3 | 73.0 | 68.7 | 68.6 | 80.2 | 88.5 | 73.2 | 79 |
| TOTAL-P | mg/L | | 12.20 | | | 13 | | | 11.7 | |
| Conductivity | mS/cm | 7.15 | 7.52 | 7.90 | 5.96 | 5.20 | 5.18 | 5.21 | 5.17 | 5.46 |
| pH | - | 7.66 | 7.61 | 7.77 | 7.69 | 7.72 | 7.70 | 7.80 | 7.82 | 7.68 |
| DO | mg/L | 2.20 | 2.17 | 0.45 | 0.51 | 3.15 | 0.40 | 0.74 | 0.53 | 0.46 |
| UV254 nm | cm-1 | 0.55 | 0.37 | 0.46 | 0.45 | 0.36 | 0.44 | 0.51 | 0.35 | 0.42 |
| DOC | mg/L | 48.70 | 32.70 | 46.80 | 46.90 | 36.04 | 40.50 | 46.70 | 41.43 | 57.90 |

| Date | Unit | 16/11/20 | 18/11/20 | 20/11/20 | 23/11/20 | 25/11/20 | 27/11/20 | 30/11/20 | 2/12/20 | 4/12/20 |
|---|---|---|---|---|---|---|---|---|---|---|
| Flow | m3/d | 140,050 | 131,050 | 130,800 | 136,350 | 138,650 | 127,850 | 177,200 | 139,300 | 171,050 |
| SS | mg/L | 1,121 | 1,074 | 1,117 | 1,131 | 1,106 | 957 | 864 | 1,055 | 836 |
| COD | mg/L | 1,554 | 1,460 | 1,367 | 1,752 | 1,536 | 1,316 | 1,273 | 1,297 | 1,187 |
| BOD5 | mg/L | 612 | 634 | 682 | 715 | 648 | 608 | 596 | 571 | 487 |
| NH4-N (Δ) | mg/L | 32.6 | 29.5 | 30.7 | 29.4 | 30.2 | 30.9 | 25.8 | 31.3 | 28.4 |
| NO3-N (Δ) | mg/L | 1.1 | 1.47 | 0.6 | 1.4 | 1.44 | 0.54 | 1.07 | 1.05 | 0.66 |
| TN | mg/L | 80.1 | 69.4 | 72.2 | 72.1 | 76.7 | 74.5 | 69.4 | 70.3 | 58 |
| TOTAL-P | mg/L | | 13.7 | | | 13.3 | | | 10.4 | |
| Conductivity | mS/cm | 5.24 | 5.56 | 5.49 | 4.86 | 4.83 | 5.23 | 5.63 | 6.40 | 6.98 |
| pH | - | 7.70 | 7.68 | 7.65 | 7.53 | 7.73 | 7.64 | 7.87 | 8.01 | 7.50 |
| DO | mg/L | 1.75 | 3.09 | 1.40 | 1.78 | 3.37 | 0.94 | 3.77 | 4.50 | 2.22 |
| UV254 nm | cm-1 | 0.40 | 0.47 | 0.45 | 0.69 | 0.48 | 0.50 | 0.85 | 0.83 | 0.41 |
| DOC | mg/L | 62.40 | 51.07 | 44.52 | 52.61 | 46.83 | 52.63 | 46.87 | 62.50 | 32.76 |

| Date | Unit | 7/12/20 | 9/12/20 | 11/12/20 | 14/12/20 | 16/12/20 | 18/12/20 | 21/12/20 |
|---|---|---|---|---|---|---|---|---|
| Flow | m3/d | 196,150 | 175,350 | 180,400 | 198,250 | 157,450 | 153,600 | 148,800 |
| SS | mg/L | 633 | 924 | 767 | 413 | 557 | 639 | 600 |
| COD | mg/L | 900 | 1,108 | 918 | 788 | 824 | 1,029 | 943 |
| BOD5 | mg/L | 422 | 472 | 368 | 335 | 342 | 391 | 436 |
| NH4-N (Δ) | mg/L | 18.5 | 40.8 | 18.1 | 17.1 | 24 | 26.9 | 29.4 |
| NO3-N (Δ) | mg/L | 0.99 | 2.3 | 1.15 | 1.25 | 1.15 | 0.92 | 2.05 |
| TN | mg/L | 40.3 | 67.8 | 45 | 35.1 | 57.3 | 56.9 | 53.6 |
| TOTAL-P | mg/L | 7.63 | 8.42 | | 5.76 | 16.8 | 9.65 | 8.55 |
| Conductivity | mS/cm | 8.07 | 15.45 | 16.52 | 16.52 | 11.20 | 8.16 | 6.85 |
| pH | - | 7.35 | 7.95 | 7.51 | 7.43 | 7.50 | 7.80 | 7.80 |
| DO | mg/L | 6.00 | 4.51 | 2.97 | 5.70 | 4.30 | 2.81 | 4.02 |

| | | | | | | | | |
|---|---|---|---|---|---|---|---|---|
| UV254 nm | cm-1 | 0.30 | 0.48 | 0.39 | 0.53 | 0.34 | 0.33 | 0.38 |
| DOC | mg/L | 30.43 | 35.40 | 31.91 | 30.30 | 34.41 | 38.10 | 39.10 |

| **Date** | **Unit** | **23/12/20** | **25/12/20** | **28/12/20** | **30/12/20** | **1/1/21** | **1/4/21** | **1/6/21** |
|---|---|---|---|---|---|---|---|---|
| Flow | m3/d | 135,180 | 141,000 | 135,150 | 135,600 | 145,850 | 175,700 | 151,470 |
| SS | mg/L | 608 | 511 | 835 | 686 | 596 | 635 | 507 |
| COD | mg/L | 979 | 868 | 1,177 | 1,023 | 1,189 | 869 | 880 |
| BOD5 | mg/L | 482 | 349 | 395 | 459 | 533 | 326 | 355 |
| NH4-N (Δ) | mg/L | 30.2 | 33.3 | 33.5 | 32.1 | 39.5 | 19.5 | 38.0 |
| NO3-N (Δ) | mg/L | 0.7 | 0.23 | 0.99 | 0.53 | 0.23 | 1.60 | 0.75 |
| TN | mg/L | 56.4 | 57.9 | 65.7 | | 70.7 | 41.5 | 88.5 |
| TOTAL-P | mg/L | 11.5 | 13.30 | 13.10 | 15.6 | 10.40 | 12.1 | 10.5 |
| Conductivity | mS/cm | 6.45 | 7 | 7.13 | 8.21 | 9 | 5.5 | 9.05 |
| pH | - | 7.78 | 7.6 | 7.77 | 7.68 | 7.4 | 7.27 | 7.64 |
| DO | mg/L | 2.90 | 2.04 | 3.3 | 3.2 | 1.66 | 5.9 | 3.12 |
| UV254 nm | cm-1 | 0.420 | 0.296 | 0.422 | 0.675 | 0.364 | 0.288 | 0.437 |
| DOC | mg/L | 47.60 | 37 | 57 | 58.3 | 43.1 | 32.6 | 38.7 |

**Table 2.** Daily numbers of COVID-19 infected people reported in the city of Thessaloniki, adjusted to specimen collection date.

| **Date** | **Reported cases** | **Date** | **Reported cases** | **Date** | **Reported cases** |
|---|---|---|---|---|---|
| 2020-10-05 | 17 | 2020-11-06 | 706 | 2020-12-08 | 237 |
| 2020-10-06 | 20 | 2020-11-07 | 696 | 2020-12-09 | 224 |
| 2020-10-07 | 23 | 2020-11-08 | 684 | 2020-12-10 | 213 |
| 2020-10-08 | 27 | 2020-11-09 | 671 | 2020-12-11 | 202 |
| 2020-10-09 | 32 | 2020-11-10 | 656 | 2020-12-12 | 190 |
| 2020-10-10 | 38 | 2020-11-11 | 639 | 2020-12-13 | 181 |
| 2020-10-11 | 45 | 2020-11-12 | 621 | 2020-12-14 | 172 |
| 2020-10-12 | 54 | 2020-11-13 | 605 | 2020-12-15 | 163 |
| 2020-10-13 | 66 | 2020-11-14 | 587 | 2020-12-16 | 155 |
| 2020-10-14 | 80 | 2020-11-15 | 569 | 2020-12-17 | 148 |
| 2020-10-15 | 97 | 2020-11-16 | 551 | 2020-12-18 | 142 |
| 2020-10-16 | 117 | 2020-11-17 | 534 | 2020-12-19 | 136 |
| 2020-10-17 | 140 | 2020-11-18 | 517 | 2020-12-20 | 130 |
| 2020-10-18 | 168 | 2020-11-19 | 499 | 2020-12-21 | 125 |
| 2020-10-19 | 199 | 2020-11-20 | 484 | 2020-12-22 | 121 |
| 2020-10-20 | 234 | 2020-11-21 | 469 | 2020-12-23 | 117 |
| 2020-10-21 | 273 | 2020-11-22 | 454 | 2020-12-24 | 113 |
| 2020-10-22 | 314 | 2020-11-23 | 438 | 2020-12-25 | 110 |
| 2020-10-23 | 358 | 2020-11-24 | 423 | 2020-12-26 | 106 |

| | | | | | |
|---|---|---|---|---|---|
| 2020-10-24 | 404 | 2020-11-25 | 410 | 2020-12-27 | 103 |
| 2020-10-25 | 449 | 2020-11-26 | 396 | 2020-12-28 | 99 |
| 2020-10-26 | 494 | 2020-11-27 | 383 | 2020-12-29 | 96 |
| 2020-10-27 | 538 | 2020-11-28 | 370 | 2020-12-30 | 93 |
| 2020-10-28 | 579 | 2020-11-29 | 356 | 2020-12-31 | 90 |
| 2020-10-29 | 616 | 2020-11-30 | 343 | 2021-01-01 | 86 |
| 2020-10-30 | 648 | 2020-12-01 | 330 | 2021-01-02 | 83 |
| 2020-10-31 | 675 | 2020-12-02 | 316 | 2021-01-03 | 80 |
| 2020-11-01 | 696 | 2020-12-03 | 303 | 2021-01-04 | 76 |
| 2020-11-02 | 715 | 2020-12-04 | 289 | 2021-01-05 | 73 |
| 2020-11-03 | 710 | 2020-12-05 | 276 | 2021-01-06 | 70 |
| 2020-11-04 | 713 | 2020-12-06 | 263 | | |
| 2020-11-05 | 711 | 2020-12-07 | 249 | | |
